# IL-6 inhibition in critically ill COVID-19 patients is associated with increased secondary infections

**DOI:** 10.1101/2020.05.15.20103531

**Authors:** Lucas M. Kimmig, David Wu, Matthew Gold, Natasha N Pettit, David Pitrak, Jeffery Mueller, Aliya N. Husain, Ece A. Mutlu, Gökhan M. Mutlu

**Affiliations:** Department of Medicine, University of Chicago, Chicago, Illinois; Section of Pulmonary and Critical Care Medicine, University of Chicago, Chicago, Illinois; Section of Infectious Diseases, University of Chicago, Chicago, Illinois; Department of Pathology, University of Chicago, Chicago, Illinois; Section of Gastroenterology and Hepatology, Rush University, Chicago, Illinois

**Keywords:** COVID-19, SARS-CoV-2, tocilizumab, cytokine release syndrome, immunosuppression

## Abstract

**Background:** Anti-inflammatory therapies such as IL-6 inhibition have been proposed for COVID-19 in a vacuum of evidence-based treatment. However, abrogating the inflammatory response in infectious diseases may impair a desired host response and predispose to secondary infections.

**Methods:** We retrospectively reviewed the medical record of critically ill COVID-19 patients during an 8-week span and compared the prevalence of secondary infection and outcomes in patients who did and did not receive tocilizumab. Additionally, we included representative histopathologic post-mortem findings from several COVID-19 cases that underwent autopsy at our institution.

**Results:** 111 patients were identified, of which 54 had received tocilizumab while 57 had not. Receiving tocilizumab was associated with a higher risk of secondary bacterial (48.1% vs. 28.1%, p=0.029 and fungal (5.6% vs. 0%, p=0.112) infections. Consistent with higher number of infections, patients who received tocilizumab had higher mortality (35.2% vs. 19.3%, p=0.020). Seven cases underwent autopsy. In 3 cases who received tocilizumab, there was evidence of pneumonia on pathology. Of the 4 cases that had not been given tocilizumab, 2 showed evidence of aspiration pneumonia and 2 exhibited diffuse alveolar damage.

**Conclusions:** Experimental therapies are currently being applied to COVID-19 outside of clinical trials. Anti-inflammatory therapies such as anti-IL-6 therapy have the potential to impair viral clearance, predispose to secondary infection, and cause harm. We seek to raise physician awareness of these issues and highlight the need to better understand the immune response in COVID-19.

## 1 INTRODUCTION

While there has been a dramatic increase in the number of clinical trials, there remains a shortage of effective therapies for COVID-19, particularly for patients who develop critical illness. The management of such patients remains largely supportive. Early reports from China suggested that an exaggerated immune response may play a role in the development of respiratory failure, shock, and multiorgan dysfunction in critically ill patients with COVID-19 (1). Similarities between the exaggerated immune response associated with COVID-19 and the cytokine release syndrome (CRS) reported in patients with CAR T-cell therapy led to the use of tocilizumab, an anti-IL-6 therapy to attenuate hyperimmune responses associated with COVID-19 (2). However, inhibition of IL-6 may also have adverse consequences. Mice lacking IL-6 response have impaired immunity against viral, bacterial and fungal pathogens (3). Humans treated with tocilizumab had higher risk of serious bacterial, skin and soft tissue infections (4-7). Lastly, IL-6 appears to play a complicated role viral clearance(8). To date, several reports detail institutional experiences with treating COVID-19 with tocilizumab.(9-13) As expected, inflammatory markers (cytokines, temperature) generally demonstrated a profound response to tocilizumab administration. However, as most reports lack a comparison group, it is difficult to ascertain whether patients clinically benefitted from inhibition of the inflammatory response. Similarly, available reports often restrict secondary infections to documented blood stream infections, which may significantly underestimate the infectious complications of anti-IL-6 therapy in the critically ill. We sought to determine the incidence of secondary infections and outcomes of patients who received tocilizumab for COVID-19 compared to those who did not in the COVID-19 intensive care unit (ICU) at our institution.

## 2 METHODS

### 2.1 Patients

We retrospectively analyzed all patients who were admitted to the COVID-19 ICU between the dates of March 1, 2020 and April 27, 2020. The study was approved by the University of Chicago Institutional Review Board. Additionally, 7 patients underwent autopsy, the histopathology of which was reviewed with our colleagues from pathology.

### 2.2 Interventions

Tocilizumab was included in our internal institutional protocol for the treatment of COVID-19, specifically for use in patients with progressive clinical deterioration and elevated inflammatory markers at the discretion of the treating team and infectious diseases consultation service. Our protocol recommended a standard dose of 400 mg of tocilizumab administered intravenously with the potential for redosing based on clinical response (e.g. oxygenation status, hemodynamic stability, inflammatory marker response). Tocilizumab was generally not considered in patients with liver function test (LFT) abnormalities, significant cytopenias, or documented ongoing infection. Similarly, patients enrolled in clinical trials (e.g. remdesivir) were not eligible to receive tocilizumab.

### 2.3 Statistical Analysis

Statistical analysis was conducted with SPSS version 26 (IBM, Armonk, NY) and R version 4 (Package Logistf). APACHE II score and Charlson comorbidity index (CCI) were not normally distributed and thus were analyzed with Mann-Whitney U-test. Count variables were analyzed with Chi-square and continuous variables were analyzed with t-tests. Multivariate regression analysis was completed in R with the outcome variable being the development of bacterial infection. The predictors (i.e., independent variables) were selected using Chi-square statistics. Variables that are statistically different between those were given tocilizumab and those who were not, were sex with the predominance of males in the tocilizumab group, history of transplant and use of immunosuppressive agents, and APACHE II scores were trending higher in the tocilizumab group (p=0.078, Mann-Whitney U test) (see table 1). There was high correlation between history of transplantation and immunosuppressive use as expected. Thus, we only included immunosuppressive use in the multivariate model. There was a trend towards a higher proportion of patients with diabetes mellitus with organ damage in those with bacterial infection and past medical history of deep venous thrombosis (DVT) or pulmonary embolism (PE). No other clinical variables were statistically different by tocilizumab use or the development of bacterial infections. Although steroid use was not statistically significantly different between those were given tocilizumab and those who were not, steroids were included in the model as they have been shown to affect COVID19 disease course and can predispose to bacterial infections. Therefore, the final model included age, sex, tocilizumab use, steroid use, diabetes with organ damage, DVT/PE, CCI, APACHE II score and immunosuppressive use. Of these, age, sex, diabetes with organ damage, CCI, and immunosuppressive use were not independently associated with the development of bacterial infections (all p>0.05). The reported odds ratios (ORs) reflect exponents of β from a Firth penalized regression model in R. Firth regression was chosen to avoid separation in the model (14).

**Table 1.**
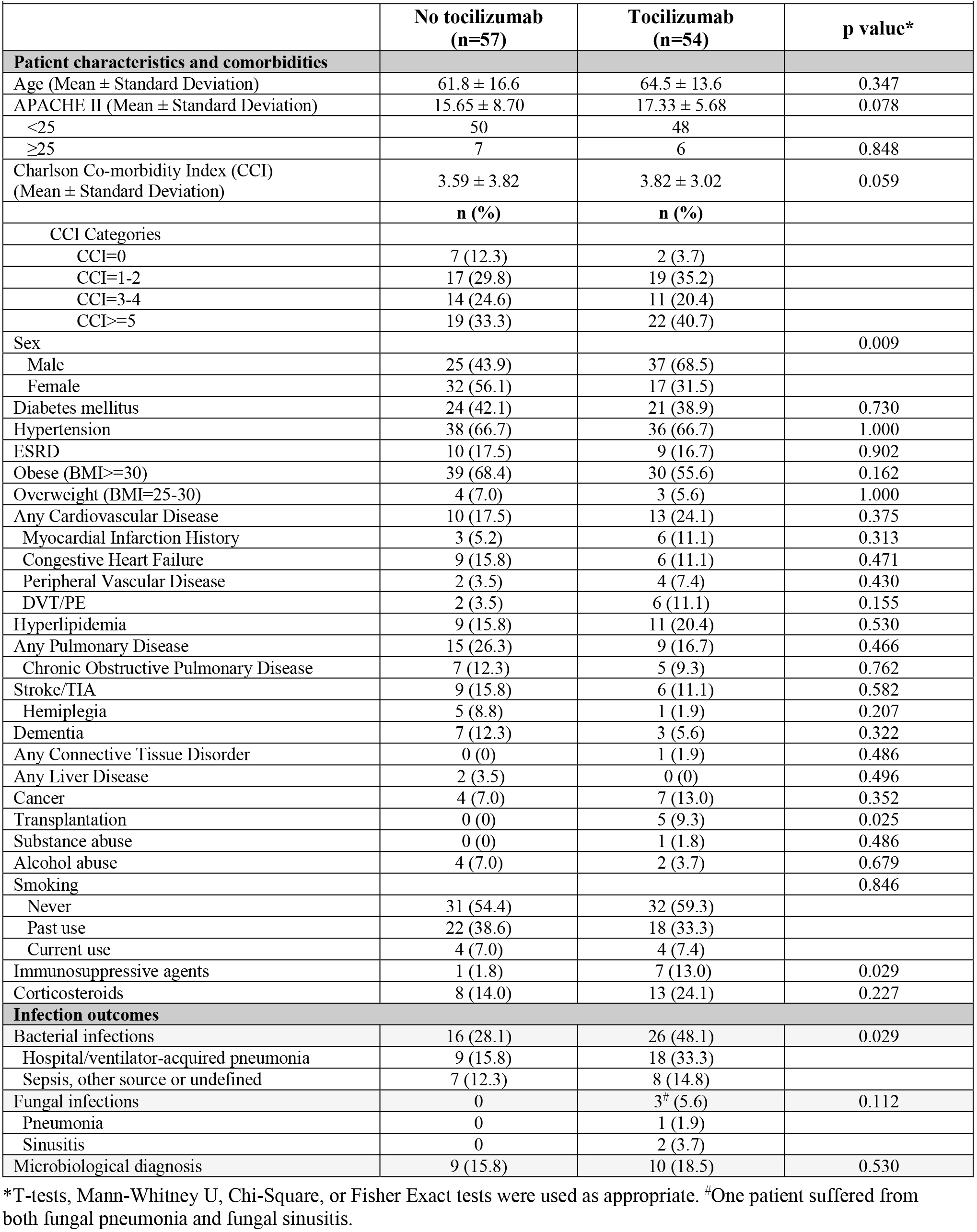
Co-morbidities and Infection outcomes.

## 3 RESULTS

### Bacterial infections are increased in critically ill patients who received tocilizumab

We identified 111 patients admitted to the COVID-19 ICU during that period. Among 54 patients who received tocilizumab, the majority (44 patients, 81%) received the standard dose of 400 mg. However, protocol deviations occurred with respect to dosing in some patients. Six patients received a total of 800 mg, one patient received 560 mg, two patients received 200 mg and one patient received 160 mg. We compared bacterial and fungal infections in those that received the drug to those that did not. Except for the male sex preponderance in tocilizumab group, there were no differences in patient baseline characteristics including CCI between the two groups (Table 1). Secondary infections were defined by positive culture data or high clinical suspicion of infection requiring the initiation of antimicrobials and documentation in the progress note. In the tocilizumab group, only infections occurring after the tocilizumab dose were counted.

Receiving tocilizumab was associated with a higher incidence of secondary bacterial infections including hospital acquired pneumonia and ventilator associated pneumonia (26 (48.1%) vs. 16 (28.1%). p=0.021). Additionally, there were three patients with fungal infections with one patient having the fungal infection in two different locations in the tocilizumab group compared with none in the non-tocilizumab group with statistical analysis showing a trend towards significance (p=0.112). Diagnosis of infection was made approximately 5 days after the administration of tocilizumab (4.9±3.0 days, median=4 days, 95% CI 3.67-6.17 days).

### Tocilizumab is independently associated with increased bacterial infections

In multivariate logistic regression model to predict bacterial infections, the following independent variables were included: age, sex, APACHE II score, CCI, immunosuppression, presence of DVT/PE, diabetes with any organ damage and use of tocilizumab and steroids. Tocilizumab use was independently positively associated with the development of bacterial infections with an odds ratio of 2.76 (95% CI 1.11-7.20) p=0.0295. APACHE II score was also independently positively associated with bacterial infections (OR: 1.08, 95% CI: 1.01-1.16, p=0.016). While steroid administration did not reach statistical significance, there was a trend towards steroids being positively associated with bacterial infections (OR: 2.76, 95% CI: 0.91-9.02, p=0.074). History of DVT/PE was negatively associated with bacterial infections (OR: 0.09, 95% CI: 0.0007–0.824). The remaining variables including age, sex, CCI, immunosuppression, and presence of diabetes with any organ damage were not independently associated with bacterial infections.

Due to well-known limitations with bacterial cultures in hospitalized and critically ill patients, we included both culture-proven and suspected bacterial infections in our analysis. While the tocilizumab group had 10 culture proven bacterial infections, non-tocilizumab group had 9 culture proven bacterial infections. This was not statistically significant. Seven patients in the non-tocilizumab and 16 patients in the tocilizumab group were treated for a bacterial infection although the cultures were negative or non-diagnostic. There was no statistical difference between the two groups in terms of culture negative bacterial infections (p=0.261).

We also compared laboratory data between two groups. In addition to the laboratory data included in the APACHE II score, we analyzed white blood cell count (WBC), percent and absolute lymphocyte count, D-dimer, C-reactive protein and ferritin, which were evaluated to determine the level of systemic inflammation as part of laboratory work up for COVID-19.

There was no difference in WBC, percent lymphocyte, D-dimer, C-reactive protein and ferritin levels between the tocilizumab and non-tocilizumab groups (Figure 1). However, mean absolute lymphocyte count was statistically lower in tocilizumab group compared to non-tocilizumab group (mean difference =0.33, p=0.02). However, there was no correlation between bacterial infections and the absolute lymphocyte count (Spearman’s rho= −0.005, p=0.959). Inclusion of the absolute lymphocyte count in the multivariate Firth regression did not change the significant variables in the model (data not shown).

**Figure 1.**
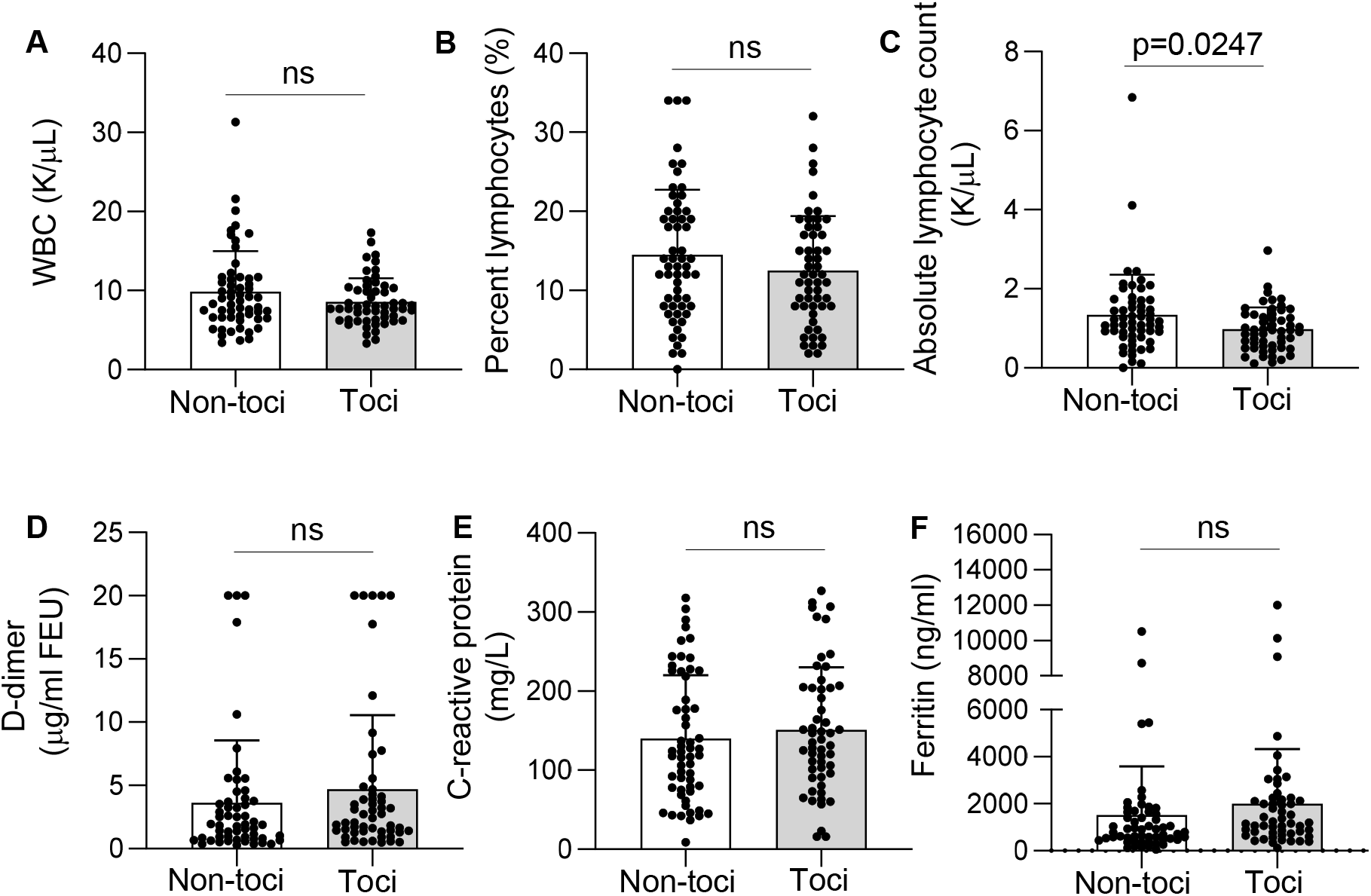
Laboratory data in tocilizumab and non-tocilizumab groups. **A**. White blood cell (WBC) count, **B**. percent lymphocyte, **C**. absolute lymphocyte count, **D**. D-dimer, **E**. C-reactive protein and F. Ferritin levels on day 1 of admission to the ICU. There was no difference between non-tocilizumab (non-toci) and tocilizumab (toci) groups in laboratory data except for absolute lymphocyte count, which was lower in the tocilizumab group. ns: not significant.

Our study did not follow outpatient status of patients treated in the ICU and not all patients had a definite outcome at the time of the study. In this preliminary analysis, compared to patients who did not receive tocilizumab, those who were prescribed tocilizumab had higher mortality (19/54 (35.2%) vs. 11/57 (19.3%, p=0.020) and a lesser number of patients were discharged home in the tocilizumab group (33.3% vs. 59.6%, tocilizumab and non-tocilizumab group, respectively) (Table 2).

**Table 2.**
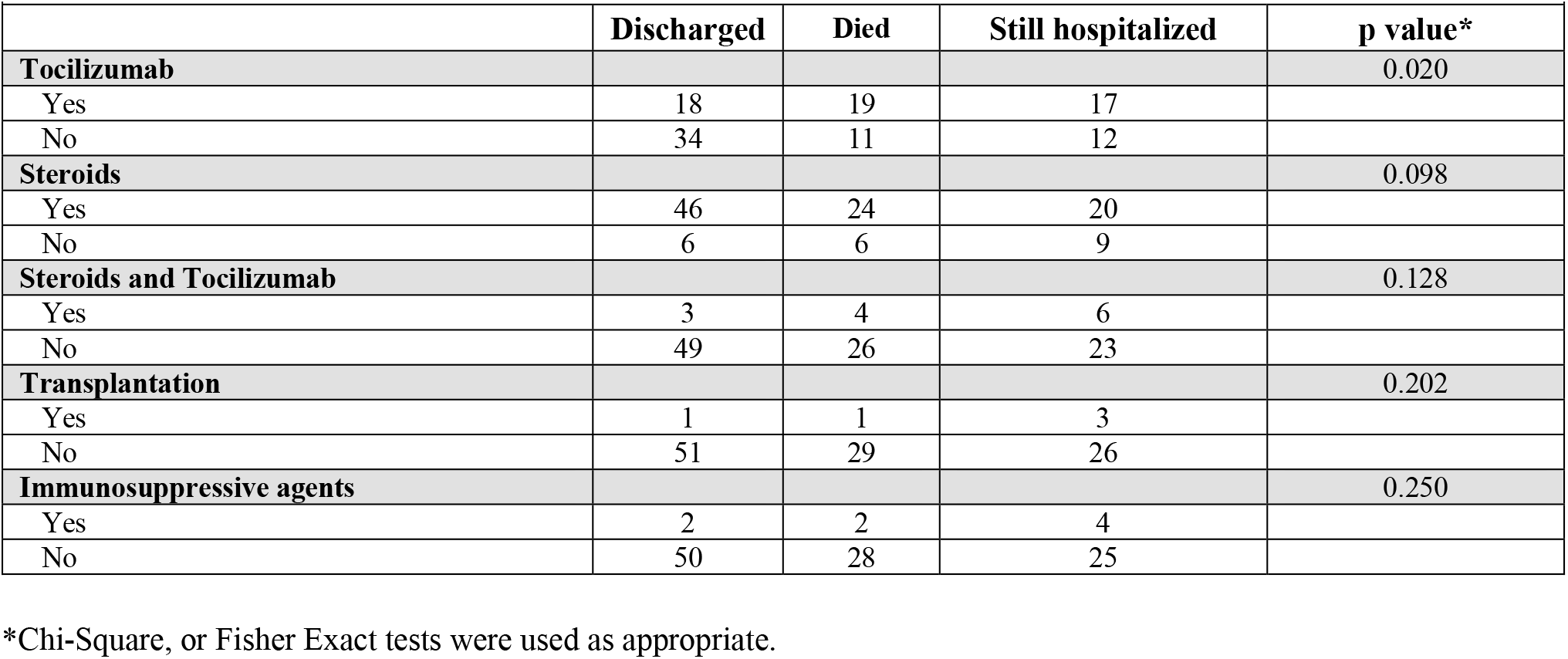
Clinical Outcomes.

IL-6 inhibition in critically ill COVID-19 patients is associated with increased secondary infections
| Table 2. Clinical Outcomes |  |  |  |  |
| --- | --- | --- | --- | --- |
|  | Discharged | Died | Still hospitalized | p value* |
| **Tocilizumab** |  |  |  | 0.020 |
| Yes | 18 | 19 | 17 |  |
| No | 34 | 11 | 12 |  |
| **Steroids** |  |  |  | 0.098 |
| Yes | 46 | 24 | 20 |  |
| No | 6 | 6 | 9 |  |
| **Steroids and Tocilizumab** |  |  |  | 0.128 |
| Yes | 3 | 4 | 6 |  |
| No | 49 | 26 | 23 |  |
| **Transplantation** |  |  |  | 0.202 |
| Yes | 1 | 1 | 3 |  |
| No | 51 | 29 | 26 |  |
| **Immunosuppressive agents** |  |  |  | 0.250 |
| Yes | 2 | 2 | 4 |  |
| No | 50 | 28 | 25 |  |

\*Chi-Square, or Fisher Exact tests were used as appropriate.

All 5 transplant patients in the study received tocilizumab. Out of these 5 patients, two were treated for a bacterial infection (one culture proven, and the other presumed based on high clinical suspicion of infection). Only one of the patients in the transplant group died. Overall transplant status did not affect outcomes. We also reviewed the administration of steroids and other immunosuppressive agents. More patients in the tocilizumab group received steroids compared to non-tocilizumab group (8 (14%) vs. 13 (24.1%), p=0.227). Administration of immunosuppressive agents, steroids alone or in combination with tocilizumab did not affect the outcomes (Table 2). After exclusion of immunosuppressed individuals from the dataset, a higher proportion of patients with bacterial infections were still observed in the tocilizumab group (16/57 vs. 24/49, p=0.034).

### Postmortem evaluation of cases

We performed post-mortem evaluation of 7 cases; 3 received tocilizumab and 4 did not. All three cases who received tocilizumab had evidence of pneumonia on pathology (Figure 2). Two of four patients who did not receive tocilizumab were nursing home residents with history of stroke and dementia and they died on the same day of admission. Their post-mortem evaluation showed evidence of aspiration pneumonia. The other two patients who did not receive tocilizumab were hospitalized for 4 and 12 days. Their lungs demonstrated only pathological changes consistent with diffuse alveolar damage without any evidence of pneumonia (Figure 2). These findings raise concerns about the use of tocilizumab to attenuate possible CRS. In particular, the occurrence of secondary bacterial infections may prolong ICU stays, and the occurrence of secondary fungal infections stands out as unusual in critical care patients without traditional risk factors (e.g. neutropenia).

**Figure 2.**
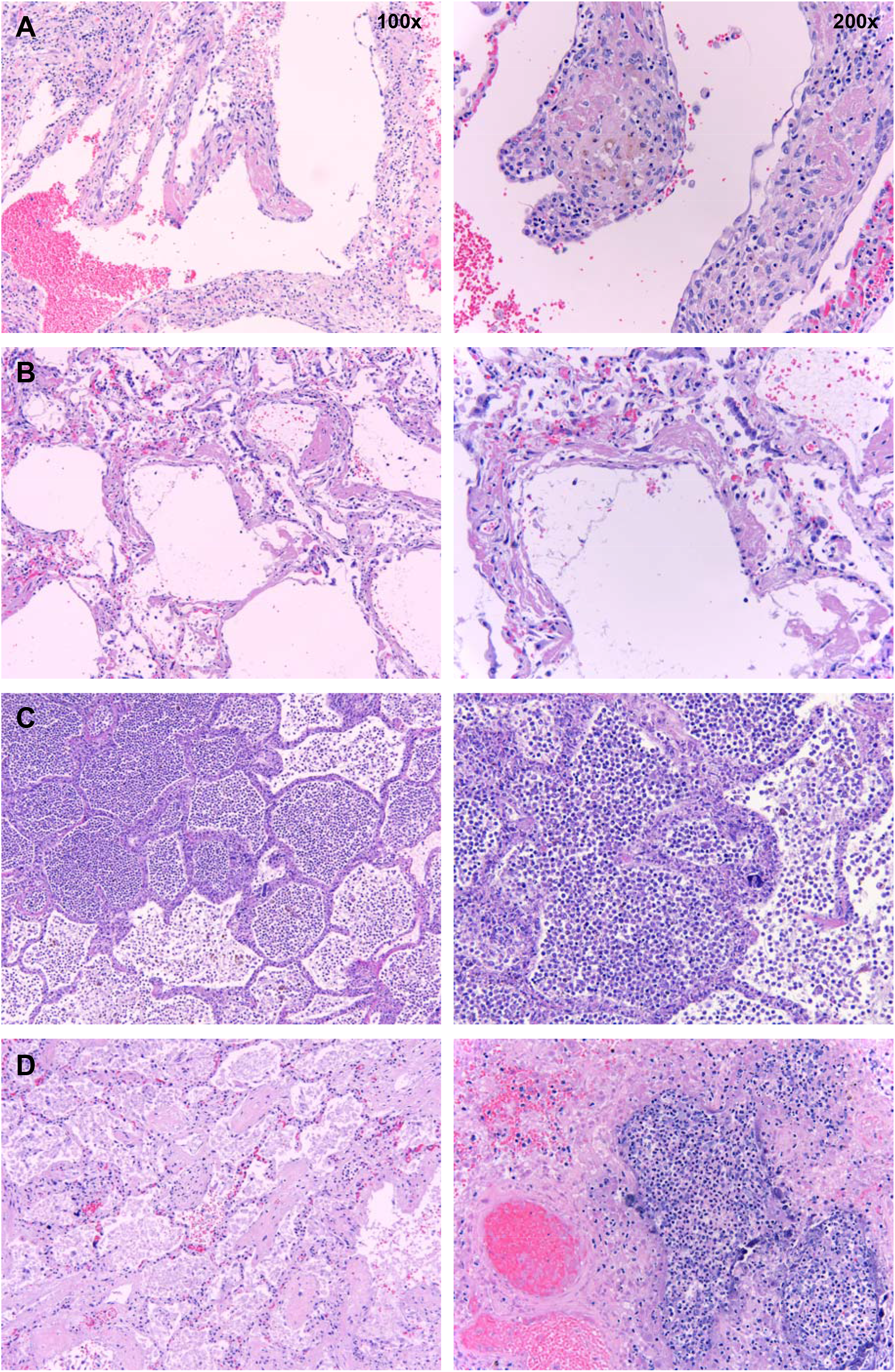
Postmortem histopathology of lungs from COVID-19 patients. Low (100x) and high power (200x) images of lungs from patients who died due to COVID-19. **A**. Organizing hyaline membranes are seen in the lung which has pre-existing emphysema (100x). Higher power shows fibrin, fibroblasts and mononuclear cells incorporated into the alveolar walls (200x). **B**. There is diffuse alveolar damage with hyaline membranes lining alveoli (100x). Higher power shows minimal inflammation with only a few mononuclear cells (200x). **C**. There is extensive intra-alveolar inflammation (neutrophils) in an otherwise normal lung (100x). On higher power, there is minimal alveolar wall thickening by inflammatory cells (also mainly neutrophils on myeloperoxidase staining and only rare lymphocytes) (200x). **D**. Majority of the sections from this case show organizing intra-alveolar fibrin (100x). Several foci of acute

## 4 DISCUSSION

Receiving tocilizumab was associated with a significantly higher rate of secondary infections and mortality at our institution. Furthermore, review of the available autopsy findings suggest that bacterial pneumonia is not uncommon in patients who die from COVID-19, particularly if they have received tocilizumab previously. These findings raise concerns about the use of anti-IL-6 therapy to attenuate a cytokine-release-like syndrome in COVID-19.

Host response to the pathogen during sepsis is a double-edged sword. Infection is generally met by a desired host inflammatory response, which can at times be vigorous. For decades, physicians have speculated whether this host response is to blame, at least in part, for the multi-organ dysfunction associated with infection, as can be seen in sepsis. In fact, the most recent definition of sepsis includes the term “dysregulated host response to infection”. According to current thought, there is orchestration of temporally distinct phases of the immune response, consisting of both initial prompt upregulation and subsequent curtailment. Dysfunctional responses may lead to overwhelming infection or persistent inflammation. Cytokines must be viewed through this lens: IL-6 may contribute to organ injury and death, but it is also central to innate immunity and microbial clearance.

At this time, it remains difficult to characterize the stage of the inflammatory response on an individual basis in real time. Similarly, many characteristics seen in severe inflammation, including, within autoregulatory limits, hypotension, may in fact represent an adaptive response. A dramatic elevation in inflammatory cytokines or other markers such as ferritin may indicate a particularly brisk host response against a more severe infection. Without proper discriminatory values that reliably identify patients in whom hyper-inflammation is the key driver of pathogenesis, treatment with anti-inflammatory therapies may be detrimental. While there may be a subset of patients who may potentially benefit from the use of tocilizumab, current evidence does not support the routine use of tocilizumab or other drugs that regulate host immune response (i.e. anti-IL1, anti-TNFα) in COVID-19 or non-COVID-19 sepsis. Evidence of efficacy for IL-6 blockade currently exists in rheumatological diseases and to manage complications of T cell engaging therapies, which are driven by a primary immune response. To date, there is no convincing evidence that immune blockade is clinically beneficial when microbial infections drive the host response. Indeed, a previous anti-cytokine strategy targeting TNFα increased mortality in septic patients (15) and was linked to increased risk of infections(16).

There are several important limitations to our study. The most important limitation is the retrospective nature and lack of randomization. Naturally, this does not allow for the establishment of causality. Because our study was retrospective, while the majority of baseline characteristics were similar, there were some that were slightly different. These include the history of transplant and use of immunosuppressive agents. Despite these limitations, patients in both groups were relatively closely matched with regards to severity of illness (APACHE II score, laboratory parameters). Patients who received tocilizumab may have developed more severe disease during the course of their ICU stay, however, our institutional policies regarding tocilizumab included important exclusion criteria, including LFT abnormalities, suspected ongoing infection, and enrollment in a clinical trial (e.g. remdesivir). In light of these limitations, it appears unlikely that the group receiving tocilizumab was excessively enriched with the sickest patients and gives credence to the descriptive power of the severity of illness captured by APACHE II scoring. While the non-tocilizumab group did not have any patients with transplantation and all patients with history of transplantation were in the tocilizumab group, history of transplantation did not affect the outcomes. Furthermore, it should be noted that the mortality data herein should be considered preliminary, because some patients were still hospitalized at the time of this analysis and all outcomes were not known.

We did not restrict our definition to rely solely on microbiologic data but defined infection as: documented infection in the medical record and antibiotic therapy initiated and continued for >2 days. This definition reflects real-world circumstances, as microbiologic identification of the causative organism is frequently not achieved in infections in the ICU, in particular for hospital-acquired and ventilator-acquired pneumonias (HAP, VAP). Furthermore, the IDSA/ATS guideline definitions of HAP and VAP do not rely on microbiologic data. Nonetheless, it is possible that our definition is too broad and may capture some patients who were treated for clinical deterioration due to COVID-19 rather than a secondary infection. However, our definition is similar to other studies where diagnoses are based on coding. Restriction to only microbiologically proven cases would likely miss a number of infections and may capture contaminants. While the numbers are small, the autopsy findings support the notion that bacterial pneumonia may be present even in absence of positive culture data.

Severe COVID-19 often elicits a strong inflammatory response with elevation in several cytokines, such as IL-6. This inflammatory phenotype shares clinical features with CAR-T cell-induced CRS and has led to the off-label use of tocilizumab for COVID-19. However, clinical similarity does not necessarily mean that changes seen in COVID-19 and CAR-T CRS are causally related and due to an overactive immune system. At least in part, the forceful immune response in COVID-19 may be adaptive and required for the anti-viral response. In our group of critically-ill COVID-19 patients, the use of tocilizumab was associated with an increase in infections. Our findings should raise physician awareness about the potential side effects of tocilizumab on pathogen clearance and development of secondary infections and additional risk vs. benefit discussions with patients and their families. It also calls for the urgent need to study all drugs including any immunosuppressive agents in randomized controlled trials to better understand their role in any hyperimmune response and on clearance of SARS-CoV-2 and other hospital-acquired pathogens, before their routine use is widely implemented.

## Data Availability

Data are available upon responsible request by contacting the corresponding author.

## 9 Author Contributions

GMM, LMK, DW, NP conceived ideas and design. GMM, LMK, DW, MG, EAM, JM, AH performed data collection. EAM, GMM, LMK, DW, MG, JM, AH, NP, DP performed data analysis and interpretation. Manuscript drafting and editing was done by GMM, LMK, DW, EAM, NP, DP.

## 11 Acknowledgements

A prior version of this manuscript was previously made available as a pre-print manuscript on *medRxiv* (doi: https://doi.org/10.1101/2020.05.15.20103531).

